# Severe COVID-19 is associated with elevated serum IgA and antiphospholipid IgA-antibodies

**DOI:** 10.1101/2020.07.21.20159244

**Authors:** Omar Hasan Ali, David Bomze, Lorenz Risch, Silvio D. Brugger, Matthias Paprotny, Myriam Weber, Sarah Thiel, Lukas Kern, Werner C. Albrich, Philipp Kohler, Christian R. Kahlert, Pietro Vernazza, Philipp K. Bühler, Reto A. Schüpbach, Alejandro Gómez-Mejia, Alexandra M. Popa, Andreas Bergthaler, Josef M. Penninger, Lukas Flatz

## Abstract

**Background:** While the pathogenesis of coronavirus disease 2019 (COVID-19) is becoming increasingly clear, there is little data on IgA response, the first line of bronchial immune defense.

**Objective:** To determine, whether COVID-19 is associated with a vigorous total IgA response and whether IgA autoantibodies are associated with complications of severe illness. Since thrombotic events are frequent in severe COVID-19 and resemble hypercoagulation of antiphospholipid syndrome (APS), our approach focused on antiphospholipid antibodies (aPL).

**Materials and methods:** In this retrospective cohort study we compared clinical data and aPL from 64 patients with COVID-19 from three independent centers (two in Switzerland, one in Liechtenstein). Samples were collected from April 9, 2020 to May 1, 2020. Total IgA and aPL were measured with FDA-approved commercially available clinical diagnostic kits.

**Results:** Clinical records of the 64 patients with COVID-19 were reviewed and divided into a cohort with mild illness (mCOVID, *n*=26 [41%]), a discovery cohort with severe illness (sdCOVD, *n*=14 [22%]) and a confirmation cohort with severe illness (scCOVID, *n*=24 [38%]). Severe illness was significantly associated with increased total IgA (sdCOVID, *P*=0.01; scCOVID, *P*<0.001). Total IgG levels were similar in both cohorts. Among aPL, both cohorts with severe illness significantly correlated with elevated anti-Cardiolipin IgA (sdCOVID and scCOVID, *P*<0.001), anti-Cardiolipin IgM (sdCOVID, *P*=0.003; scCOVID, *P*<0.001), and anti-Beta2 Glycoprotein-1 IgA (sdCOVID and scCOVID, *P*<0.001). Systemic lupus erythematosus was excluded from all patients as a potential confounder of APS.

**Conclusions:** Higher total IgA and IgA-aPL were consistently associated with severe illness. These novel data strongly suggest that a vigorous antiviral IgA-response triggered in the bronchial mucosa induces systemic autoimmunity.

## INTRODUCTION

The novel coronavirus disease 2019 (COVID-19) caused by severe acute respiratory syndrome coronavirus 2 (SARS-CoV-2) has become a global pandemic with wide-ranging health and socio-economic implications. After SARS and the Middle East Respiratory Syndrome, it represents the third known spillover of a severe coronavirus-associated disease from animals to humans in the last twenty years (1-3). SARS-CoV-2 enters human cells by attachment to and subsequent internalization of Angiotensin-converting enzyme 2 receptors that are highly expressed by type-II pneumocytes in the deep bronchial system (4), where IgA immunoglobulins produced in the bronchial-associated lymphoid tissue (BALT) are the main line of humoral defense (5). Indeed, specific IgA against the SARS-CoV-2 spike protein have been shown to appear early in infected patients (6). However, despite the important role of IgA in mucosal immunity, the rate of total IgA generated by that response and its role in COVID-19 severity remains unexplored.

Autopsies have shown acute respiratory distress syndrome (ARDS) and sepsis to be the most common complications in critically-ill COVID-19 patients (7). A large case series from Northern Italy that assessed lung histologies of deceased COVID-19 patients presents consistent diffuse alveolar damage and necrosis of pneumocytes (8). A distinctive factor for COVID-19 was a marked presence of diffuse thrombosis of the peripheral small vessels. This is in line with reports of frequent thromboembolisms of patients with severe COVID-19 that occur despite the prophylactic in-hospital use of low weight molecular heparins (9, 10). While the reasons remain unclear, a recent report describes a case series of severe COVID-19 patients with stroke and elevated levels of antiphospholipid (aPL) antibodies compatible with antiphospholipid syndrome (APS) (11). APS is an acquired autoimmune disease that is mediated by autoantibodies directed against phospholipid-binding proteins that leads to hypercoagulability. The most common trigger factor of APS is systemic lupus erythematosus (SLE), followed by lung infections with mycobacteria or viruses (12). These cases suggest that COVID-19 leads to virally triggered APS in severe COVID-19 patients. To our knowledge, complete aPL profiling in mild and severe cases of COVID-19 has never been undertaken. The aim of this study was to explore, whether patients with severe COVID-19 have elevated total IgA as an immediate immune response and aPL compatible with APS.

## METHODS

This multicenter cohort study was conducted at the three following tertiary care hospitals: the Landesspital Liechtenstein (LLS) in Vaduz, Liechtenstein, the Kantonsspital St. Gallen (KSSG) in St. Gallen, Switzerland, and the University Hospital Zurich (USZ) in Zurich, Switzerland. The collection of patient data and blood samples were approved by the respective local ethics committees of the participating study centers (Project-IDs 2020-00676, 2020-00821, and 2020-00646). All participants agreed to the hospitals’ general consent policies allowing further use of clinical data and biologic material (LLS and KSSG) or signed an informed consent (USZ). Where signing of informed consent was not possible due to severe illness and to prevent surface contamination, consent was sought verbally or from the next of kin, which had been approved by the local ethics committee.

### Collection of data

Samples of serum or plasma anticoagulated with lithium heparinate (LHP) were collected from April 9, 2020, to May 1, 2020. We included patients with SARS-CoV-2 infection that was confirmed either by real-time reverse transcriptase-polymerase chain reaction (RT-PCR) or serology. RT-PCR was performed and interpreted in strict adherence to World Health Organization guidelines (13). SARS-CoV-2 antibodies were analyzed by two different antibody tests: a lateral flow immunochromatographic assay (LFIA, gold nanoparticle-based, SGIT flex Covid 19, Sugentech, South Korea) and an electro-chemiluminescence immunoassay (ECLIA, Elecsys Anti-SARS-CoV-2, Roche Diagnostics International Ltd, Switzerland). Seropositivity was defined as a positive IgG result in the LFIA that was confirmed in the ECLIA.

Patients were categorized into the following cohorts: 1. mild illness without requirement of hospitalization (mCOVID), 2. a discovery cohort of patients with severe illness (sdCOVID), and 3. an independent confirmation cohort of patients with severe illness (scCOVID). Mild illness was defined as uncomplicated upper respiratory tract infection with unspecific symptoms or uncomplicated pneumonia, while severe illness for the sdCOVID and scCOVID cohorts required hospitalization and included severe pneumonia, ARDS, sepsis and septic shock (13).

### Immunoglobulins and aPL testing

aPL antibodies were determined by fluorescence enzyme immunoassay on a Phadia 250 analyzer (Thermo Fisher Diagnostics AG, Steinhausen, Switzerland) using EliATM Cardiolipin as well as EliATM Beta 2-Glycoprotein 1 assays for IgG, IgA, and IgM isotoypes (all Thermo Fisher Diagnostics AG, Steinhausen, Switzerland). Total IgA and IgG were determined on a Cobas c501 analyzer (Roche Diagnostics, Rotkreuz, Switzerland) with a nephelometric assay (IgA-2, Tina-quant IgA Gen.2, and IgGu2, Tina-quant IgG Gen.2, Roche Diagnostics, Rotkreuz, Switzerland). IgG against SS-A/Ro, SS-B/La, dsDNA, Sm, chromatin and RNP were simultaneously determined by bead-based suspension array principle on a Bioplex 2200 System (Biorad Laboratories, Cressier, Switzerland). Coefficients of variations, as determined by commercially available control materials were 4.5% for total IgA, <2.0% for total IgG, 4.6% for aPL antibodies, and 3.7% for lupus-antibodies. All measurements were performed in the same laboratory (Labormedizinisches Zentrum Dr. Risch, Vaduz, Liechtenstein).

### Statistical methods

Point estimates of autoantibody levels were described using the mean and the standard error of the mean. Differences in autoantibody levels between cohorts were assessed using the nonparametric Mann-Whitney test. All *P*-values were adjusted for multiple hypothesis testing using the false discovery rate (FDR) method. Statistical significance was defined at the level of FDR <0.05. All analyses were performed using R software, version 3.5.0 (R Project for Statistical Computing, Vienna, Austria). Furthermore, we generated hierarchical clustering (spearman clustering distance and a complete clustering method) on the median centered values of antibody levels. Missing values were replaced with the respective mean value across the remaining patients.

## RESULTS

### Patient characteristics

We collected a total of 64 serum or LHP samples from patients with SARS-CoV-2 infection (one sample per patient). The mean age of all patients was 62 years (IQR, 46-74) and 32 (60%) were male. 26 (41%) only had mild symptoms without the need for hospitalization and were allotted to the mCOVID cohort. The average age among mildly symptomatic patients was 57 (IQR, 45-63) years and 9 (35%) were male. 38 (59%) patients had severe COVID-19 and required 8 hospitalization: the sdCOVID cohort included 14 (22%) and the scCOVID 24 (38%) patients. Their mean age was 70 (IQR, 58-76) years and they were predominantly male (28 [74%]). 59 (92%) of 64 patients had been confirmed SARS-CoV-2 positive via real-time polymerase chain reaction (RT-PCR), which included all patients from the sdCOVID and scCOVID cohorts, as well as 19 patients from the mCOVID cohort. The remaining 7 (11%) patients from the mCOVID cohort were confirmed by serology.

4/26 (15%) patients in the mCOVID cohort had co-morbidities predisposing to severe COVID-19, of which the most frequent was hypertension (3/26 [12%]). In contrast, 26/28 (93%) severely ill patients had at least one co-morbidity, of which the most common were hypertension (26/38 [68%]), cardiovascular disease (25/38 [66%]) and diabetes mellitus (13/38 [34%]). Of note, 13/38 (34%) of patients with severe COVID-19 developed thromboses during hospitalization, while none of the mCOVID patients reported thronboses. Patient characteristics are listed in **Table 1**.

**Table 1.** Overview of patient characteristics.

| Patient characteristics | Mild illness<br>(n=26) | Severe illness,<br>Discovery cohort<br>(n=14) | Severe illness,<br>Confirmation cohort<br>(n=24) |
| --- | --- | --- | --- |
| Sex - No. (%) |  |  |  |
| Male | 9 (35) | 8 (57) | 20 (83) |
| Female | 17 (65) | 6 (43) | 4 (17) |
| Median age (IQR) - yr | 57 (45-63) | 64 (56-70) | 73 (64-80) |
| Comorbidities* - No. (%) |  |  |  |
| No | 22 (100) | 2 (14) | 0 (0) |
| Yes | 4 (15) | 12 (86) | 24 (100) |
| Hypertension | 3 (12) | 10 (71) | 16 (67) |
| Cardiovascular disease | 0 (0) | 6 (43) | 19 (79) |
| Cerebrovascular disease | 0 (0) | 0 (0) | 6 (25) |
| Diabetes | 1 (4) | 5 (36) | 8 (33) |
| Liver disease | 0 (0) | 0 (0) | 5 (21) |
| Cancer | 0 (0) | 0 (0) | 9 (38) |
| Kidney disease | 0 (0) | 5 (36) | 7 (29) |
| Thromboembolisms, No. (%) |  |  |  |
| Yes | 0 (0) | 4 (29) | 9 (38) |
| No | 26 (100) | 10 (71) | 15 (63) |
| Immunosuppression, No. (%)† |  |  |  |
| Yes | 0 (0) | 5 (36) | 5 (21) |
| No | 26 (100) | 9 (64) | 19 (79) |

IQR: interquartile range; No.: number; yr: years. \* Comorbidities only consider diagnoses that are known risk factors for developing severe COVID-19, as listed. † Immunosuppression is defined as systemically applied prednisone $\geq 7.5$ mg/day (or equivalent), other systemic immunosuppressive drugs, such as calcineurin inhibitors, and chemotherapy.
Percentages adding to >100 are due to number rounding.

### Antibody results

Severely ill COVID-19 patients had significantly higher total IgA titers compared to mCOVID patients (sdCOVID, mean 2.94 g/l, SD ±0.46, *P*=0.01; scCOVID mean 3.04 g/l, SD ±0.19, *P*<0.001), but not higher total IgG (sdCOVID mean 7.69 g/l, SD ±0.55, *P*=0.09; scCOVID not measured). They also had significantly higher anti-Cardiolipin IgA (sdCOVID mean 6.38 U/ml, SD ±0.96, *P*<0.001; scCOVID mean 4.86 U/ml, SD ±0.84, *P*<0.001), anti-Beta2 Glycoprotein-1 IgA (sdCOVID mean 8.50 U/ml, SD ±3.86, *P*<0.001; scCOVID mean 4.71 U/ml, SD±2.17, *P*<0.001), and Cardiolipin IgM (sdCOVID mean 4.01 U/ml, SD ±0.88, *P*=0.003; scCOVID mean 10.35 U/ml, SD ±5.48, *P*<0.001), as shown in **Figure 1**. With two other aPL antibodies we found a significant difference only in the sdCOVID but not the scCOVID cohort: anti-Cardiolipin IgG (sdCOVID mean 8.23 U/ml, SD ±4.02, *P*=0.02; scCOVID mean 2.42, SD ±0.54, *P*=0.09) and anti-Beta2Glycoprotein-1 IgG (sdCOVID mean 1.57 U/ml, SD ±0.23, *P*=0.002; scCOVID mean 1.58 U/ml, SD ±0.85, *P*=0.15). No significant difference was found among anti-Beta2 Glycoprotein-1 IgM among the cohorts (sdCOVID mean 1.07 U/ml, SD ±0.25, *P*=0.16; scCOVID mean 2.00 U/ml, SD ±0.72, *P*=0.16), as shown in **Figure 2**. Hierarchical clustering analysis further indicated common elevation of antibody titers among patients with mild and severe COVID-19 (see **Figure 3**).

**Figure 1.**
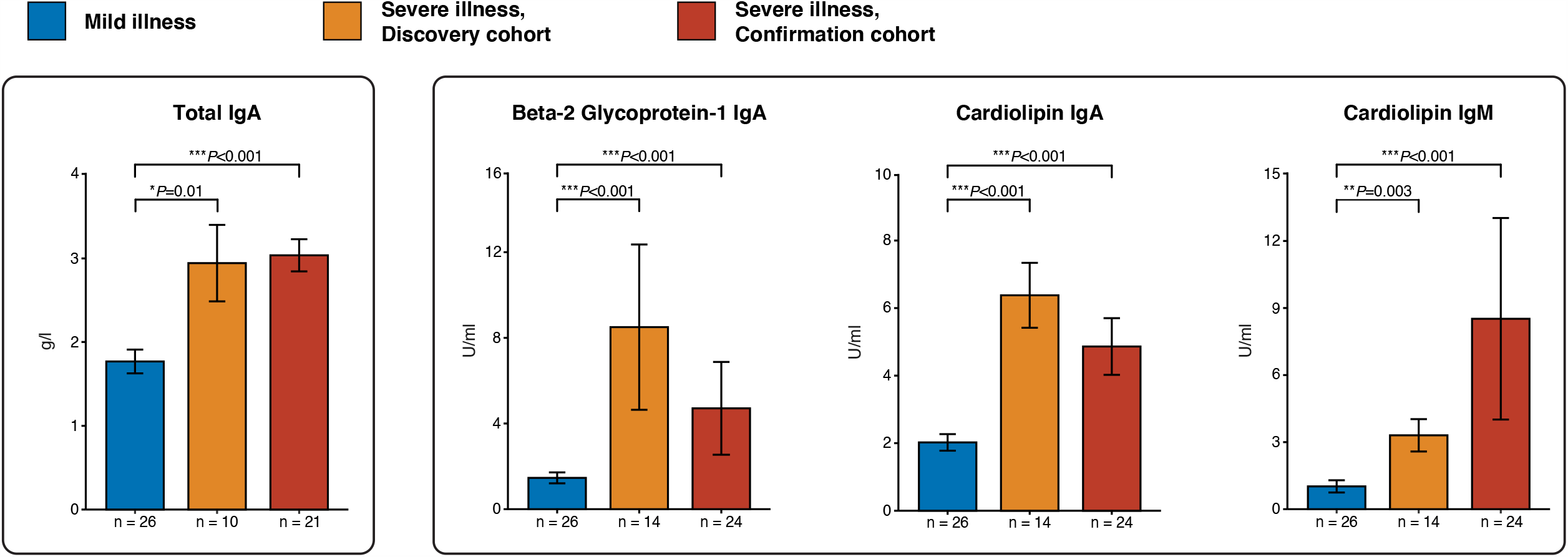
Antibodies with a significant association with severe COVID-19 in both cohorts (discovery and confirmation). Plot titles indicate the respective protein targets and type of immunoglobulin. Y-axes reflect measured results and units. X-axes display patient counts.

**Figure 2.**
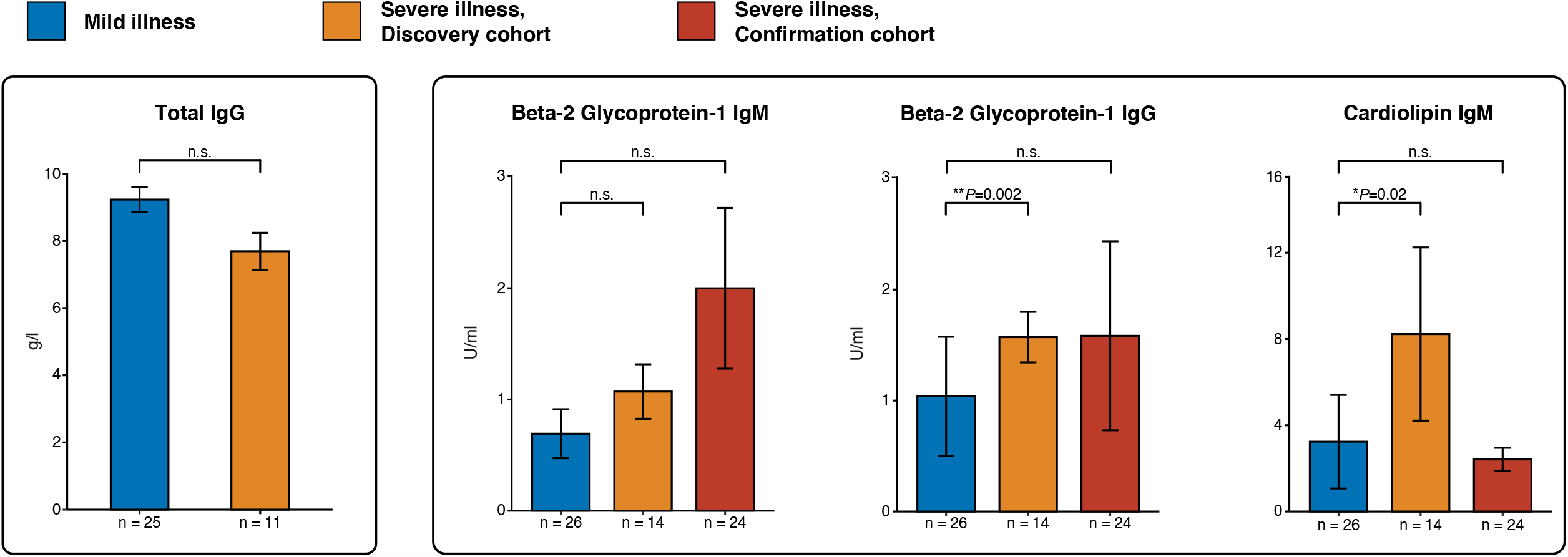
Antibodies with no or only partial significance with severe COVID-19. Plot titles indicate the respective protein targets and type of immunoglobulin. Y-axes reflect measured results and units. X-axes display patient counts.

**Figure 3.**
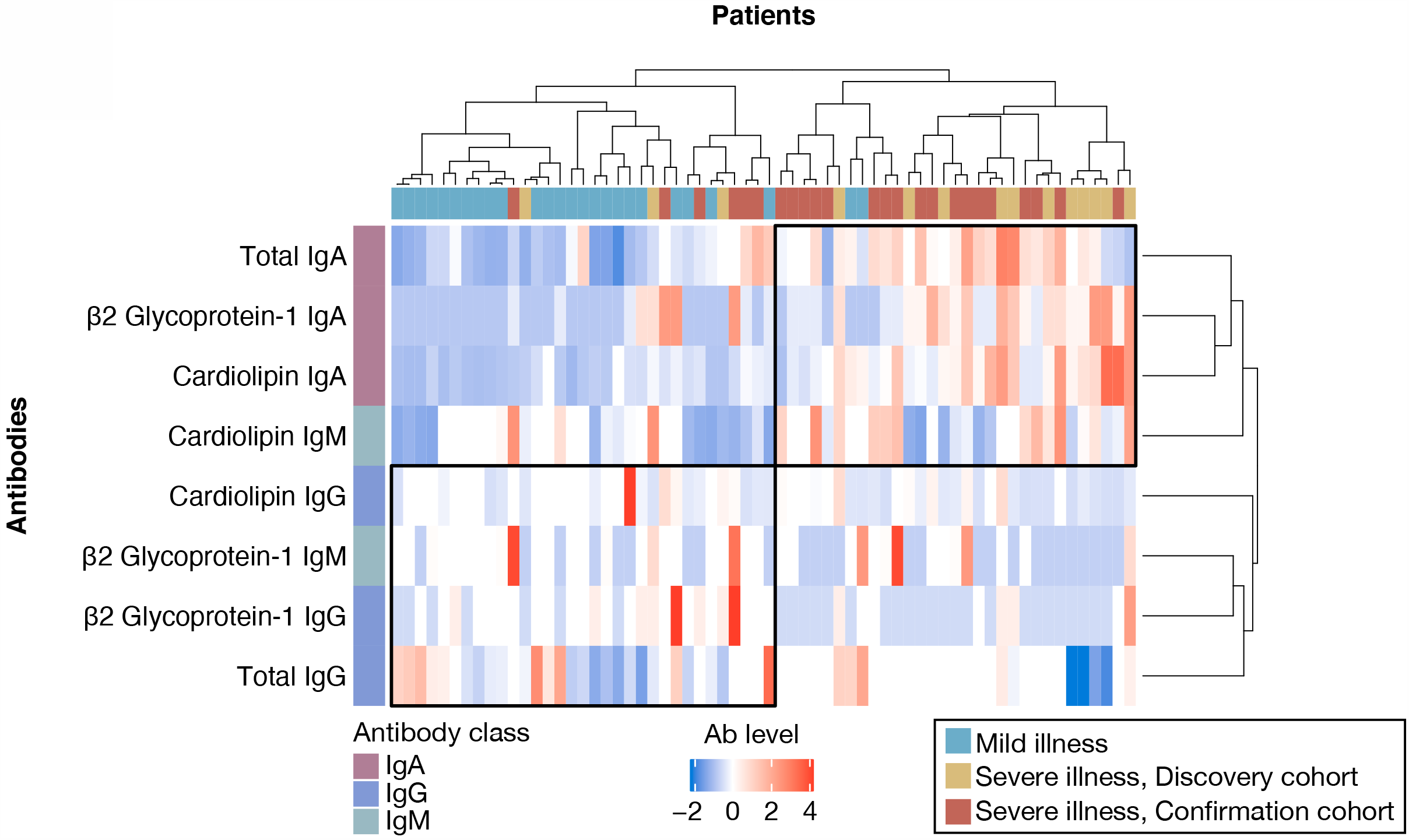
Heatmap of the hierarchical clustering of antibody levels across patients. Clustering depicts common shifts of antibody levels among patients with mild illness and severe COVID-19. Rows display antibody levels and columns list patients. All antibody levels are median centered and are displayed as fold relative compared to the median, with higher titers displayed in red and lower titers in blue. For missing results, the mean value of the respective antibody/immunoglobulin was used. The scale bar (bottom center) denotes the x-fold levels.

Since APS is commonly triggered by SLE serology screening was performed for all patients, as described in the methods section. One patient from the scCOVID cohort had elevated anti-La IgG (7.4 U/mL, normal < 1 U/mL) but no other lupus-specific antibodies. None of the other patients were positive for any SLE-associated antibodies.

## DISCUSSION

In this study we measured total IgA and IgG, as well as aPL in COVID-19 patients of similar age and compared the results of mildly ill with severely ill patients. For severely ill patients we established a discovery and investigation cohort, allowing eliminating potential center-specific confounders. Our novel finding shows a marked elevation of total IgA that is significantly associated with severe COVID-19, which to our knowledge has not been reported before. There is no significant association with total IgG. These data are in line with our hypothesis, that a strong, IgA-driven immune response emerges from the BALT when SARS-CoV-2 affects the deeper respiratory system. In line with literature, about a third of severely ill patients developed thromboses (14). To explain thromboembolisms, we found that total IgA and aPL IgA antibodies were significantly associated with severe illness. This correlation could neither be seen for total IgG, nor for aPL IgG antibodies. It is likely that a deep bronchial viral infection leads to a stronger immune response in the BALT than an upper respiratory infection, which may explain the high level of IgA in severely ill patients compared to patients with mild disease. (15) While an association of aPL and severe COVID-19 has been established, which strongly supports our findings (16), the elevation of total IgA together with IgA-aPL draws the missing link between immune response and hypercoagulation through induction of IgA-dominated APS.

APS is most commonly triggered by SLE (12). To eliminate this potential confounder, we conducted a lupus serology of all patients in our cohort that showed no indications for SLE in our cohorts (17). Another potential trigger of APS is infection including pneumonia, that may trigger a molecular mimicry mechanism (18-20). In the case of COVID-19 such as mechanism is likely mediated by pulmonary surfactant, as it is rich in phospholipid-binding proteins (21). Surfactant is produced by type-II pneumocytes, that express high levels of ACE-2 receptors and are effectively destroyed during severe COVID-19 (8, 14). Pneumocyte necrosis leads to surfactant leakage and the peptide commonalities between SARS-CoV-2 and surfactant proteins may expose phospholipid protein structures for mimicry. Indeed, a high coverage of peptide commonalities has recently been demonstrated by Kanduc D. et al., who found that almost half of the immunoreactive epitopes on the spike glycoprotein of SARS-CoV-2 share pentapeptides on human surfactant-related proteins (22). In practice, preliminary results of the COVID-19 therapy (RECOVERY) trial demonstrate a significant benefit of dexamethasone therapy for severely ill patients (23), suggesting the role of immune exacerbation in COVID-19 related deaths. The effects of aPL could be further enhanced by toll-like receptor-4, which is upregulated in SARS (24) and has been shown to enhance hypercoagulation (25).

Importantly, our data further supports a link between IgA and Kawasaki disease-like multisystem inflammatory syndrome in children (MIS-C) (26), since Kawasaki disease can be triggered by respiratory viruses and severe forms are associated with organ deposits of IgA-producing plasma cells (27, 28).

Potential limitations of the study are its cohort sizes and retrospective design. Nevertheless, the inclusion of a discovery and an independent confirmation cohort from different health care centers is as well as similar age among cohorts are major strengths of the study. These data contribute to our understanding of the potential role of autoimmunity in severe COVID-19.

In conclusion, we present a novel significant association between severe COVID-19, elevated total IgA and IgA-aPL. These findings suggest that autoantibodies may be causally linked to COVID-19 severity and thrombosis. We hypothesize that COVID-19 is a potent inductor of autoimmunity and recommend further studies with larger cohorts and mechanistic exploration to validate these findings.

## Data Availability

The authors confirm that the data supporting the findings of this study are available within the article.

## ACKNOWLEDGEMENTS

We thank Dorothea Hillmann and Francesca Ferrara (both from Labormedizinisches Zentrum Dr. Risch) for laboratory analyses without receiving compensation.

